# Suicide Deaths during the Stay-at-Home Advisory in Massachusetts

**DOI:** 10.1101/2020.10.20.20215343

**Authors:** Jeremy S. Faust, Sejal B. Shah, Chengan Du, Shu-Xia Li, Zhenqiu Lin, Harlan M. Krumholz

## Abstract

Many believe that shelter-in-place or stay-at-home policies might cause an increase in so-called deaths of despair. While increases in psychiatric stressors during the COVID-19 pandemic are anticipated, whether suicide rates changed during stay-at-home periods has not been described.

This was an observational cohort study that assembled suicide death data for persons aged 10 years or older from the Massachusetts Department of Health Registry of Vital Records and Statistics from January 2015 through May 2020. Using autoregressive integrated moving average (ARIMA) and seasonal ARIMA to analyze suicide deaths in Massachusetts, we compared the observed number of suicide deaths in Massachusetts during the stay-at-home period (March through May, 2020) in Massachusetts to the projected number of expected deaths. To be conservative, we also accounted for the deaths still pending final cause determination

The incident rate for suicide deaths in Massachusetts was 0.67 per 100,000 person-month (95% CI 0.56-0.79) versus 0.81 per 100,000 person-month (95% CI 0.69-0.94) during the 2019 corresponding period (incident rate ratio of 0.83; 95% CI 0.66-1.03). The addition of the 57 deaths pending cause determination occurring from March through May 2020 and the 33 cases still pending determination from the 2019 corresponding period did not change these findings.

The observed number of suicide deaths during the stay-at-home period did not deviate from ARIMA projected expectations using either preliminary data or an alternate scenario in which deaths pending investigation (exceeding the average remaining number of deaths still pending investigation which occurred during the corresponding 2015-2019 period) were ascribed to suicide. Decedent age and sex demographics were unchanged during the pandemic period compared to 2015-2019.

The stable rates of suicide deaths during the stay-at-home advisory in Massachusetts parallel findings following ecological disasters. As the pandemic persists, uncertainty about its scope and economic impact may increase. However, our data are reassuring that an increase in suicide deaths in Massachusetts during the stay-at-home advisory period did not occur.

## Introduction

Many policymakers believe that shelter-in-place or stay-at-home policies might cause an increase in “deaths of despair”. While increases in psychiatric stressors during the COVID-19 pandemic are anticipated and have been reported, whether suicide rates changed during stay-at-home periods has not been described.^1,2^

## Methods

We assembled suicide death data for persons aged ≥10 years from the Massachusetts Department of Health Registry of Vital Records and Statistics from January 2015 through May 2020.

We used autoregressive integrated moving average (ARIMA) and seasonal ARIMA models to analyze suicide deaths in Massachusetts using yearly population as the covariate. We used the Akaike information criterion (AIC) to select the best model. We plotted suicide deaths during each month of 2020 for which adequate data exist (January through May). To be conservative, we plotted an alternative scenario in which the number of deaths pending investigation by the state medical examiner exceeding 2015-2019 corresponding monthly averages were ascribed to suicide. Incident rates and rate ratios for the pandemic period (March through May) and the corresponding period in 2019 were determined. STROBE guidelines were followed. This study was not subject to institutional review board approval because it used public data.

## Results

During the pandemic period, the incident rate for suicide deaths in Massachusetts was 0.67 per 100,000 person-month (95% CI 0.56-0.79) versus 0.81 per 100,000 person-month (95% CI 0.69-0.94) during the 2019 corresponding period (incident rate ratio of 0.83; 95% CI 0.66-1.03). Because data for 2019 and 2020 are “preliminary,” a sensitivity analysis including all deaths still pending final cause adjudication as of 10/14/2020 was performed. The addition of the 57 deaths pending cause determination occurring from March through May 2020 and the 33 cases still pending determination from the 2019 corresponding period did not change these findings. The conservative assumption that all pending investigations for March-May were suicides yielded an incident rate of 0.94 deaths (95% CI 0.81-1.08) per 100,000 person-month for 2020 and 0.97 deaths (95% CI 0.84-1.11) for 2019) per 100,000 person-month (incident rate ratio 0.97; 95% CI 0.80-1.18).

The number of suicide deaths during the stay-at-home period did not deviate from projected expectations using either preliminary data or an alternate scenario in which deaths pending investigation (exceeding the average remaining number of deaths still pending investigation which occurred during the corresponding 2015-2019 period) were ascribed to suicide (Figure). Decedent age and sex demographics were unchanged during the pandemic period, compared to 2015-2019. (Table).

## Discussion

The stable rates of suicide deaths during the stay-at-home advisory in Massachusetts parallel findings following ecological disasters.^3^ Early in the outbreak, social distancing-created stressors may have been offset by a sense of shared purpose in “flattening the curve.” There were efforts at bolstering connections through video platforms, anticipation about governmental support including unemployment benefits and stimulus aid, and mental health awareness campaigns about the risks of isolation, loneliness, and despair that could accompany the public health imperative of physical distancing.

One limitation to this study is its reliance on cause of death adjudication. However, unlike other causes of death, every suicide is investigated by a medical examiner, rendering resulting death certificates more reliable than many other causes.

As the pandemic persists, uncertainty about its scope and economic impact may increase.^4^ Suicide risk often increases with rising unemployment and related strains, access to firearms, substance use, and interpersonal violence.^5^ Decompensation of the seriously mentally ill may also contribute to related morbidity and mortality.

However, our data are reassuring that an increase in suicide deaths in Massachusetts during the stay-at-home advisory did not occur. Moving forward, effective prevention efforts will require comprehensive attention to the full spectrum of mental health services.

## Data Availability

Data Statement and Author Contributions:
Dr. Faust had full access to all of the data in the study and takes responsibility for the integrity of the data and the accuracy of the data analysis.
Concept and design: Faust, Shah, Lin, Krumholz
Acquisition, analysis, or interpretation of data: Faust, Du, Li, Lin
Drafting of the manuscript: Faust, Shah, Du, Krumholz
Critical revision of the manuscript for important intellectual content: All authors.
Statistical analysis: Faust, Du, Li, Lin
Supervision: Faust, Lin, Krumholz

## Data Statement and Author Contributions

Dr. Faust had full access to all of the data in the study and takes responsibility for the integrity of the data and the accuracy of the data analysis.

*Concept and design:* Faust, Shah, Lin, Krumholz

*Acquisition, analysis, or interpretation of data:* Faust, Du, Li, Lin Drafting of the manuscript: Faust, Shah, Du, Krumholz

*Critical revision of the manuscript for important intellectual content:* All authors.

*Statistical analysis:* Faust, Du, Li, Lin

*Supervision:* Faust, Lin, Krumholz

## Funding Statements

Faust: None.

Shah: None.

Du: None.

Li: None.

Lin: Dr Lin reported working under contract with the Centers for Medicare & Medicaid Services.

Krumholz: In the past three years, Harlan Krumholz received expenses and/or personal fees from UnitedHealth, IBM Watson Health, Element Science, Aetna, Facebook, the Siegfried and Jensen Law Firm, Arnold and Porter Law Firm, Martin/Baughman Law Firm, F-Prime, and the National Center for Cardiovascular Diseases in Beijing. He is an owner of Refactor Health and HugoHealth, and had grants and/or contracts from the Centers for Medicare & Medicaid Services, Medtronic, the U.S. Food and Drug Administration, Johnson & Johnson, and the Shenzhen Center for Health Information.

---

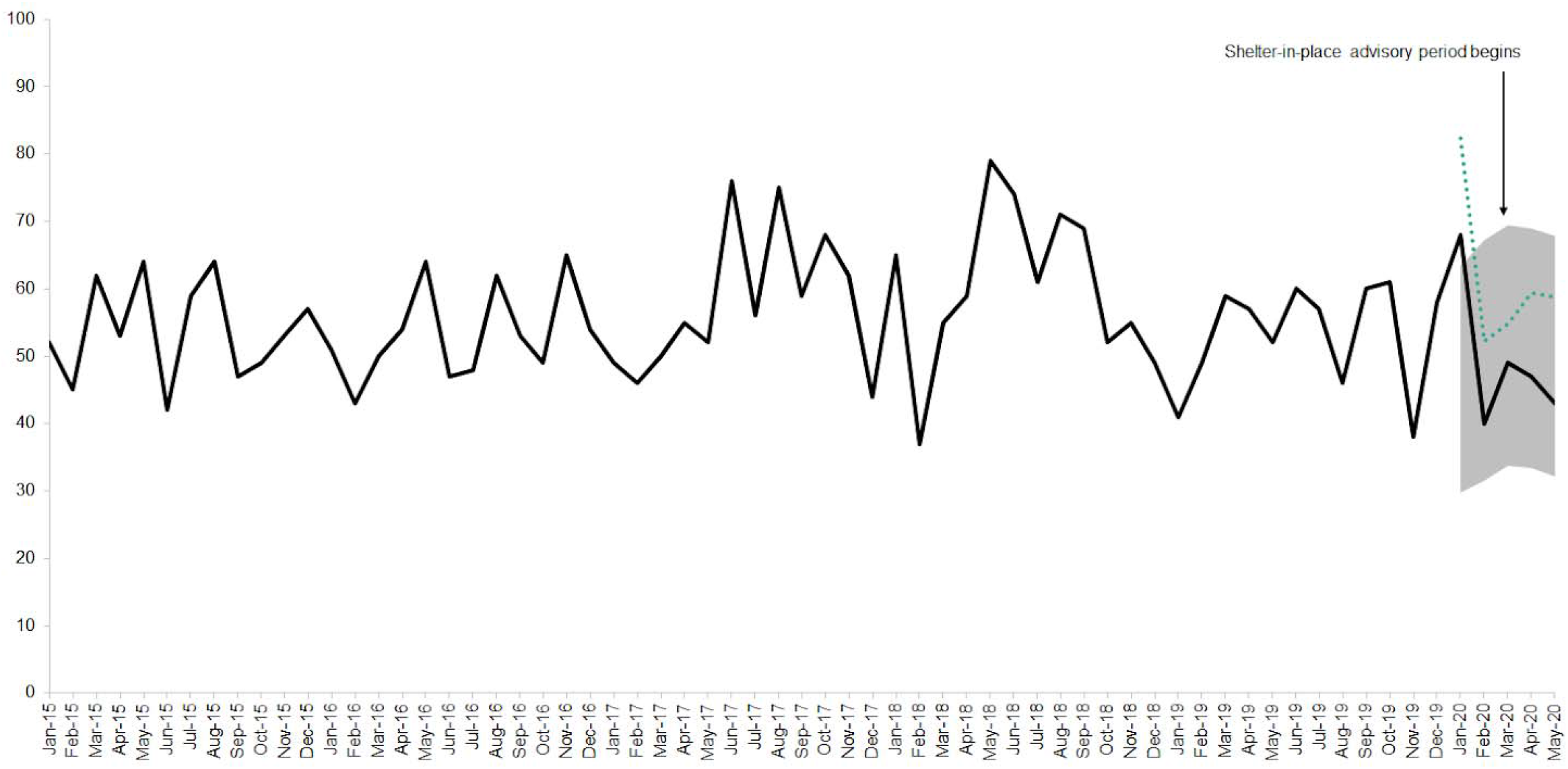
<u>Suicide deaths in Massachusetts from January 2015 through May 2020.</u> *Black line:* raw suicide death counts. *Green dots:* raw suicide deaths plus deaths pending investigation by the state medical examiner that are in excess of monthly averages of active pending investigations during the corresponding months from 2015-2019. *Grey shaded area:* the projected range of suicide deaths expected to occur during 2020 (seasonal adjusted model).

|  | **Percent of**<br>**deaths, female** | **Percent of**<br>**deaths, male** | **Mean age,**<br>**female** | **Mean**<br>**age, male** | **Mean**<br>**age, all** |
| --- | --- | --- | --- | --- | --- |
| **2015** | 24.4 | 75.6 | 47.6 | 47.5 | 47.6 |
| **2016** | 23.1 | 76.9 | 47.5 | 44.7 | 45.3 |
| **2017** | 24.2 | 75.8 | 49.7 | 44.1 | 45.5 |
| **2018** | 23.3 | 76.7 | 46.3 | 46.3 | 46.3 |
| **2019** | 22.3 | 77.7 | 44.4 | 50 | 48.76 |
| **2020** | 24.5 | 75.5 | 44.7 | 49.9 | 48.6 |

## Notes

### Funding Statement

There was no funding for this project.

### Author Declarations

This study was not subject to institutional review board approval because it used public data.

## References

1. Reger MA, Stanley IH, Joiner TE. Suicide Mortality and Coronavirus Disease 2019—A Perfect Storm? JAMA Psychiatry. Published online April 10, 2020. doi:10.1001/jamapsychiatry.2020.1060

2. Moutier C. Suicide Prevention in the COVID-19 Era: Transforming Threat Into Opportunity. JAMA Psychiatry. Published online October 16, 2020. doi:10.1001/jamapsychiatry.2020.3746

3. Morganstein JC, Ursano RJ. Ecological Disasters and Mental Health: Causes, Consequences, and Interventions. Front Psychiatry. 2020;11:1. doi:10.3389/fpsyt.2020.00001

4. Phillips JA, Nugent CN. Suicide and the Great Recession of 2007-2009: the role of economic factors in the 50 U.S. states. Soc Sci Med. 2014;116:22–31. doi:10.1016/j.socscimed.2014.06.015

5. Gunnell D, Appleby L, Arensman E, et al. Suicide risk and prevention during the COVID-19 pandemic. The Lancet Psychiatry. 2020;7(6):468–471. doi:10.1016/S2215-0366(20)30171-1

